# Statistical Methods for Evaluating Contrived Sample Functional Characterization Studies for Next-generation Sequencing-based Qualitative Assays

**DOI:** 10.1101/2025.11.08.25339816

**Authors:** Li Guan, Paul Wenz, Kristen Meier

## Abstract

Evaluation of new clinical assays involves numerous studies using patient samples to verify performance. Due to limited patient sample availability, supplementary surrogate samples may be needed. To evaluate functional comparability between surrogate and patient samples, a contrived sample functional characterization (CSFC) study has been recommended. Alongside this, a statistical analysis framework has been proposed by Godsey et al. for NGS-based assays to quantify differences in overall probit regressions and C*X* levels (estimated concentrations of the measurand at which an assay declares a sample positive X% of the time). However, detailed statistical methods for implementing this proposed framework remain unestablished. This work evaluates statistical methods for framework implementation. Using simulated data, we compared two approaches for evaluating differences between sample types using centered and non-centered probit models: test for interaction and bootstrap for differences in probit regressions. While both approaches showed similar performance in detecting overall differences, the centered model more frequently detected true differences than the non-centered model. Additionally, when evaluating differences in C*X* levels, in most scenarios using C95 alone was often insufficient in detecting true differences in overall regressions whereas testing all five C*X*s showed elevated type □ error rate. Therefore, while both approaches have strengths and limitations, we recommend using the interaction term approach with the centered model to evaluate the differences in overall probit regression as the primary approach and applying the bootstrap approach when C*X* levels are of interest, when pooling surrogate and patient samples is inappropriate, or when verification of model-based findings is desired.

## 1. Background

With a proven ability to simultaneously scan a person’s entire genome, next-generation sequencing (NGS) technology has been increasingly utilized as a platform to develop companion diagnostic (CDx) assays in oncology. Cancer, driven by changes to certain genes, is the second-leading cause of death in the US when considering all age groups combined (1). Cancer CDx assays help identify patients who may benefit from a specific drug or therapy or experience serious adverse events. For instance, the US Food and Drug Administration (FDA) approved the TruSight™ Oncology (TSO) Comprehensive test (2), an in vitro diagnostic assay, as a CDx for detecting NTRK1/2*β* fusion in patients with solid tumors, to determine eligibility for the treatment with VITRAKVI^®^ (larotrectinib) (2). This assay also detects RET fusions in patients with non-small lung cancer, helping to identify those who may benefit from using RETEVMO^®^ (selpercatinib) (2).

To ensure NGS-based assays provide accurate and reliable results, robust analytical validation studies are necessary to demonstrate how well an assay can detect disease-causing DNA or RNA variants in a patient sample. Conducting a suite of well-designed analytical validation studies requires a rich inventory of patient samples collected from the assay intended use population. For example, a single-site precision study recommended by the Clinical and Laboratory Standards Institute (CLSI) EP5 (3) requires 80 aliquots each from several patient samples. However, in practice obtaining an adequate number of patient samples with sufficient volume can be challenging and sometimes impossible due to a variety of reasons including the low prevalence of targeted biomarkers and the invasive procedure of obtaining patient samples. NGS based assays interrogate multiple biomarkers simultaneously, which exacerbates the challenge of obtaining a sufficient number of patient samples since the samples need to represent all the different biomarkers and potentially multiple cancer types.

To overcome the difficulty posed by the limited availability of the full range of patient samples, surrogate samples may be used to supplement patient samples in analytical validation studies. When considering the use of surrogate samples, it is crucial to thoroughly assess qualities of the surrogate samples that may impact assay performance following the hierarchical approach outlined in CSLI EP39 (4). For NGS-based assays, regulatory bodies (5,6), and BloodPAC Analytical Variables Working Group (7) have recommended that developers evaluate the functional comparability between surrogate and patient samples through a contrived sample functional characterization (CSFC) study. CLSI does not provide specific guidelines for the study design and analysis approach for CSFC studies whereas Godsey et al. (7) provides some guidance. An overview of the recommended CSFC study according to Godsey et al. (7) with an illustrative example of an NGS-based assay is provided.

Consider an NGS-based qualitative reporting assay that detects disease-causing DNA variants (point mutations) in a particular gene in a patient sample as an example to illustrate the recommended CSFC study design. The assay quantifies the variant allele frequency (VF) of the disease-causing variant for each position in the gene as an internal continuous response, and ultimately reports a binary outcome, positive or negative, to represent the presence or absence of each mutation. The goal of a CSFC study is to assess whether surrogate and patient samples are functionally comparable across different dilutions levels near the assay’s variant detection cutoff. For assays where the detection cutoff is the limit of blank (LOB), the CSFC study evaluates samples near the limit of detection (LOD); for assays where the detection threshold is higher than LOB, the CSFC study evaluates samples near the C95. The C*X* value (e.g. C95) is the VF level where the assay declares a sample to be positive X% (e.g. 95%) of the time.

To conduct a CSFC study, the assay developers will start with a patient sample and a surrogate sample containing the same mutational variant at the same VF at the top level and create serial dilutions. The serial dilution is recommended to include at least five levels around or below C95, for example, C95, C75, C50, C25, and C5, with at least 20 replicates for each level per sample. Next, the NGS-based assay will be conducted in parallel between the surrogate and patient sample replicates. For the purposes of comparing sample types, test performance is characterized by modelling the binary outcomes as a function of VF, commonly through probit regression. Conceptually, if the two sample types are functionally equivalent, then for each sample type we will expect to see similar probit regression lines, stemming from similar model parameters (intercept and slope), predicting similar hit rates (test positive rates) at the same VF levels (or inversely predicting similar C*X* values). Therefore, Godsey et al. (7) recommend comparing the differences between sample types by reporting point estimates and two-sided 95% confidence intervals (CIs) for the differences in model parameters and key C*X* values.

To implement this recommended statistical evaluation, methodology for making comparisons between sample types while quantifying uncertainty and statistical significance is paramount to robust CSFC analysis. While it is straightforward to obtain the point estimates for differences in the model parameters and estimated C*X* levels between sample types, obtaining CIs for the differences remains challenging and ambiguous. In this paper, using simulated data mimicking actual data observed from NGS-based qualitative assays, we explored two statistical approaches using probit regression to detect differences between surrogate and patient samples: modelling with an interaction term (interaction term approach) and constructing CIs from a bootstrap distribution (bootstrap approach). While this study is motivated by NGS-based assays, the findings and recommendations may also be applicable to other binary assays, such as those based on Polymerase Chain Reaction (PCR) techniques.

## 2. Methods

### 2.1. Simulating Data to Mimic an NGS-based CSFC Study

In this simulated study example motivated by real-world data, we consider an NGS-based assay intended to detect small DNA variants in specific genes in a tumor tissue. The assay calculates the percent mutational VF as an intermediate output and ultimately reports whether a variant is detected (test positive) or not detected (test negative). The CSFC study includes six dilution levels to achieve VF levels around and below C95.

VFs and test results (positive or negative) for a targeted small DNA variant were simulated for six dilution levels corresponding to VF levels (expressed in decimal units between 0 and 1) 0.02, 0.04, 0.05, 0.06, 0.085, and 0.12. For each sample type and each level, the VF *X*_*ij*_ at level i (i=1,2,…,6) for a replicate j was randomly generated from a normal distribution *X*_*ij*_ *∼ N (µ*_*i*_, sd = 0.005), where *µ*_*i*_ was equal to 0.02, 0.04, 0.05, 0.06, 0.085, and 0.12 for the six dilution levels, respectively. The standard deviation of 0.005 was set constant across levels. The outcome *Y*_*ij*_ was generated from a Bernoulli distribution with parameter 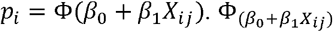 is the standard normal cumulative distribution function for *β*_0_ *+ β*_1_*X*_*ij*_. CLSI EP17 (8) recommends using 40 replicates (20 replicates per reagent lot) per sample per dilution level when determining LOD with the probit approach. Accordingly, 40 replicates were simulated per dilution level for each sample (j=1, 2, …, 40).

Five scenarios were simulated, each with a pair of surrogate and patient samples (Table 1), representing a variety of characteristic differences between sample types. The theoretical probit regressions generated using the parameters (*β*_0_ and *β*_1_) in Table 1 are displayed in Figure S1. In Scenario 1, data were simulated with the same *β*_0_ and *β*_1_ and hence the difference between the surrogate and patient sample types was only driven by random variations. In the other four scenarios, data were simulated with different *β*_0_ and/or *β*_1_.

**Table 1.**
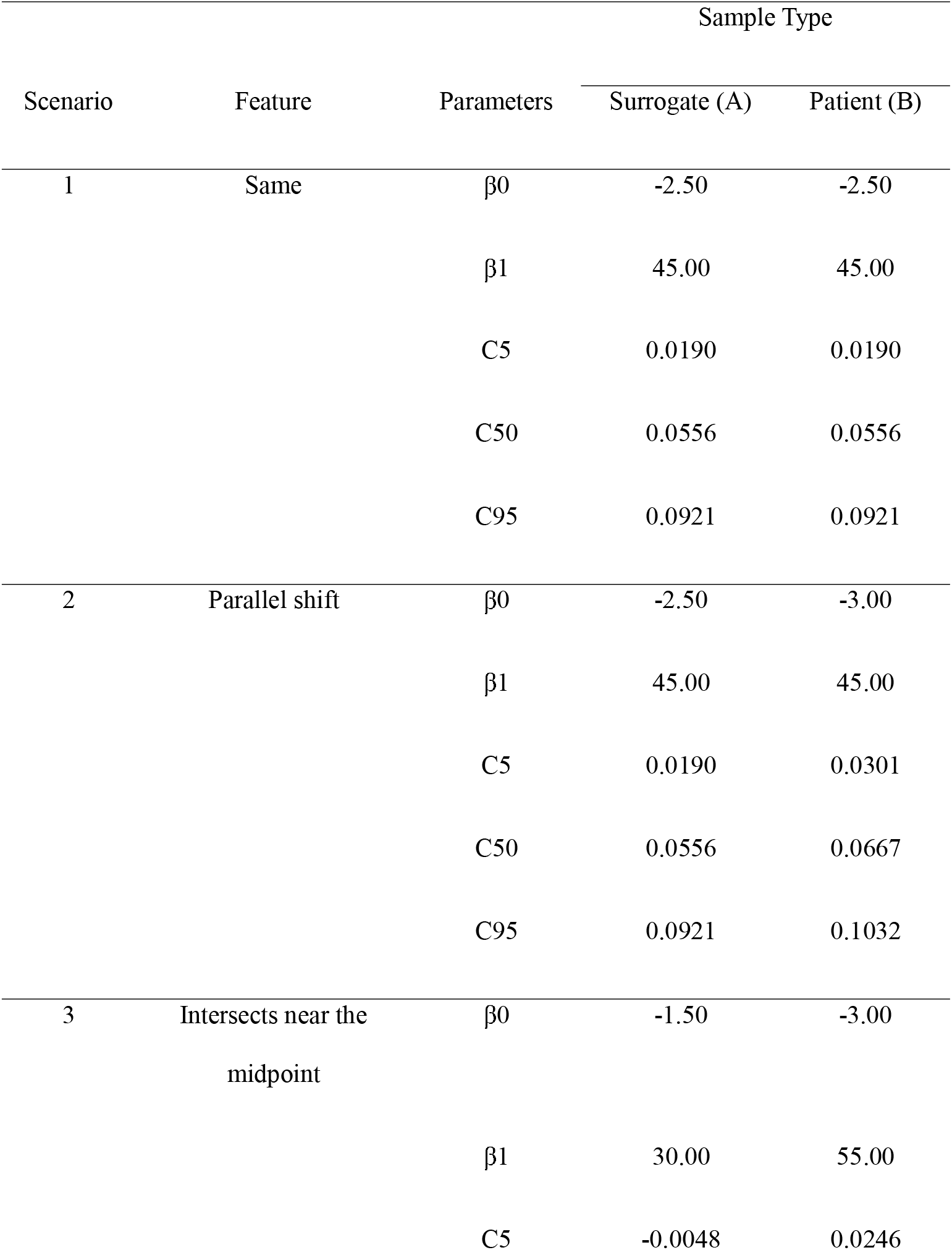
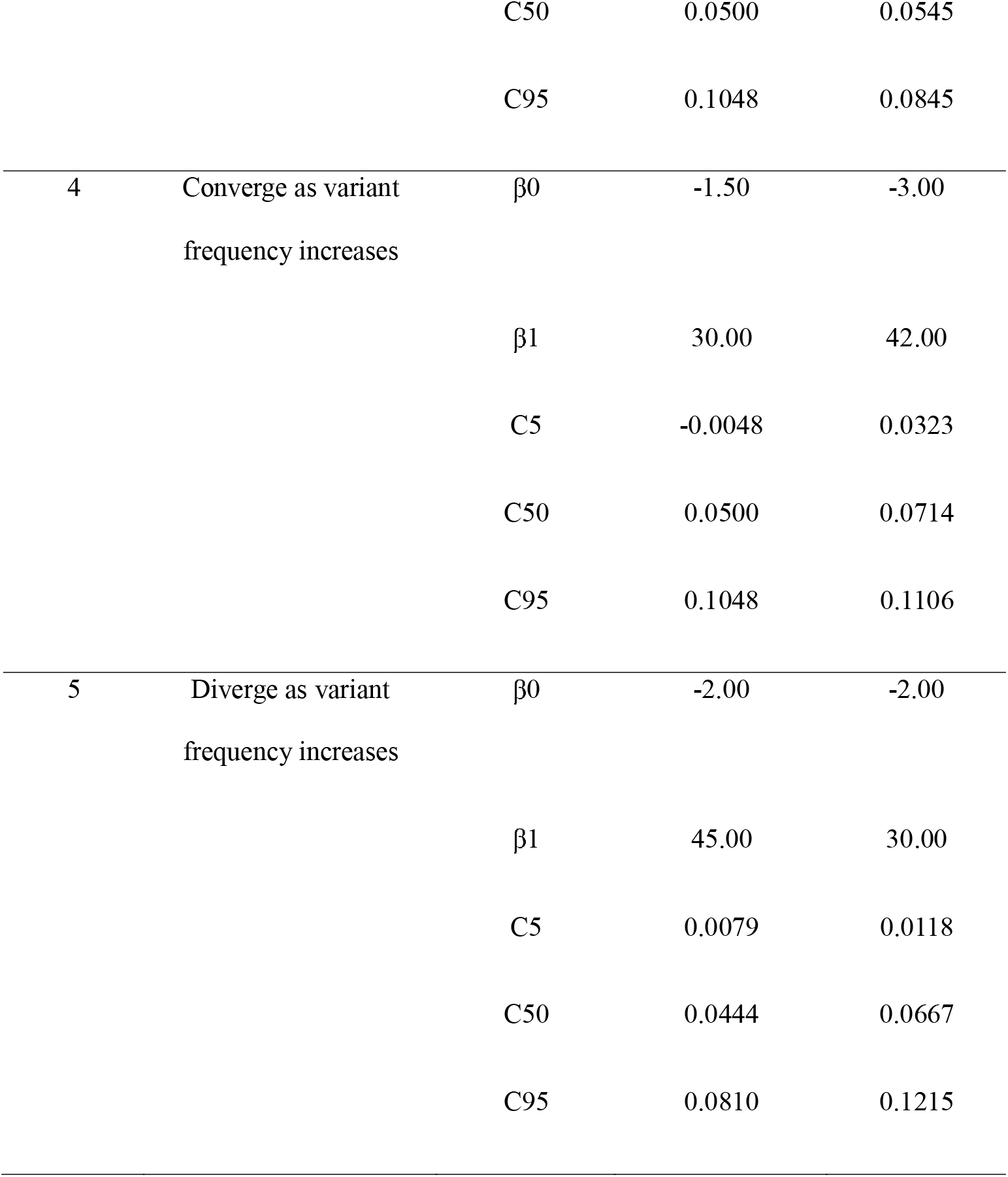
Probit regression model parameters and derived theoretical C5, C50 and C95 levels used in five simulation scenarios.

| Scenario | Feature | Parameters | Sample Type |  |
| --- | --- | --- | --- | --- |
|  |  |  | Surrogate (A) | Patient (B) |
| 1 | Same | $\beta_0$ | -2.50 | -2.50 |
| | | $\beta_1$ | 45.00 | 45.00 |
|  |  | C5 | 0.0190 | 0.0190 |
|  |  | C50 | 0.0556 | 0.0556 |
|  |  | C95 | 0.0921 | 0.0921 |
| 2 | Parallel shift | $\beta_0$ | -2.50 | -3.00 |
| | | $\beta_1$ | 45.00 | 45.00 |
|  |  | C5 | 0.0190 | 0.0301 |
|  |  | C50 | 0.0556 | 0.0667 |
|  |  | C95 | 0.0921 | 0.1032 |
| 3 | Intersects near the<br>midpoint | $\beta_0$ | -1.50 | -3.00 |
| | | $\beta_1$ | 30.00 | 55.00 |
|  |  | C5 | -0.0048 | 0.0246 |
|  |  | C50 | 0.0500 | 0.0545 |
|  |  | C95 | 0.1048 | 0.0845 |
| 4 | Converge as variant<br>frequency increases | $\beta_0$ | -1.50 | -3.00 |
| | | $\beta_1$ | 30.00 | 42.00 |
|  |  | C5 | -0.0048 | 0.0323 |
|  |  | C50 | 0.0500 | 0.0714 |
|  |  | C95 | 0.1048 | 0.1106 |
| 5 | Diverge as variant<br>frequency increases | $\beta_0$ | -2.00 | -2.00 |
| | | $\beta_1$ | 45.00 | 30.00 |
|  |  | C5 | 0.0079 | 0.0118 |
|  |  | C50 | 0.0444 | 0.0667 |
|  |  | C95 | 0.0810 | 0.1215 |

### 2.2. Fitting Probit Regressions Within Individual Samples

With the simulated CSFC study data of VF *X*_*ij*_ and binary outcomes *Y*_*ij*_ from a pair of surrogate and patient samples, statistical analyses following Godsey et al. (7) were performed. First, for each sample, the mean VF among all the replicates for a given level i was computed and denoted as 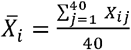, and the hit rate for level i was computed and denoted as 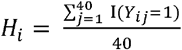. Next, for each sample, a probit regression model was fit to the binary outcomes *Y*_*ij*_ versus mean VFs across the six levels following US FDA recommendations and recognized standards (7,8) as shown in Model 1 below.

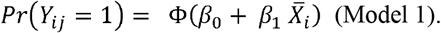

*β*_0_ and *β*_1_ and their corresponding 95% CIs were obtained from probit regressions for surrogate and patient samples, separately. In addition, the estimated hit rate at a given mean VF was calculated using forward prediction and its 95% Wald CI was computed using R Studio version 4.1.0; the estimated VF at a given hit rate was calculated with an inverse prediction and its 95% CI was computed using the Fieller’s theorem with JMP software (version 17.0.0; SAS Institute Inc, Cary, North Carolina).

The estimated intercept of Model 1 represents the probability of detecting the mutational variant when VF is equal to zero. Alternatively, a centered model (Model 2) can be fit by centering the predictor 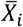, as shown below.

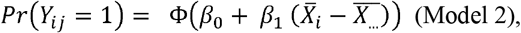

where 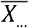 is the mean of the VFs across replicates spanning the six dilution levels of the two samples. The estimated intercept of Model 2 represents the probability of detecting the mutational variant when VF is equal to the mean VF across replicates spanning the six levels of the two samples.

### 2.3 Obtaining Point Estimates and CIs for the Differences in Model Parameters and in Predicted CX Levels Between Samples

Once the probit model (Model 1 or 2) was fit to individual samples, the point estimates of the differences in the probit intercepts and slopes between sample types were easily obtained by subtraction. Next, the C*X* levels were estimated from the inverse prediction based on the fitted probit regressions, and then the point estimates of the differences in predicted C*X* levels between sample types were obtained in the same way. However, obtaining the CIs for these differences is challenging; to construct these CIs, both the interaction term and bootstrap approaches were utilized.

#### 2.3.1 Interaction Term Approach

To test for differences in the probit intercepts and slopes between surrogate and patient samples, data from the two sample types were pooled together and a probit model with an interaction term for VF (dilution) level by sample type (Model 3) was fit using R Studio version 4.1.0.

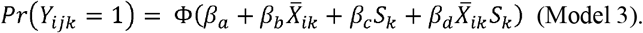

In Model 3, *Y*_*ijk*_ indicates the binary outcomes at level i for replicate j from sample 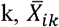 indicates the mean of VFs at level i from sample k across the 40 replicates, and the *S*_*k*_ is an indicator variable with the value of 1 denoting sample k is a patient sample. The parameters *β*_*C*_ and *β*_*d*_ are the differences in intercepts and slopes between probit regressions for surrogate and patient samples, respectively. Differences were considered statistically different when their two-sided 95% CI did not include 0. While the CIs for *β*_*C*_ and *β*_*d*_ of Model 3 are readily obtained from the output of the Fit Model function in JMP software (version 17.0.0; SAS Institute Inc, Cary, North Carolina), the CIs for the differences in the estimated C*X* levels cannot be directly derived from Model 3.

Similar to how both centered and non-centered models can be applied in probit regressions for individual samples (section 2.2), the centered model can also be used when applying the interaction term approach with the pooled data, as shown below.

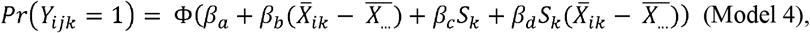

where 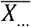 is the mean of the VFs across replicates spanning the six dilution levels of the two samples.

#### 2.3.2 Bootstrap Approach

An alternative approach based on bootstrap was used to construct CIs for the predicted C*X* levels, and it was also used as another way to construct CIs for differences in the intercepts and slopes of probit regressions. For each bootstrap run, sampling with replacement was performed among the 40 replicates within each dilution level and sample. Next, using the bootstrap data, the same analysis described in 2.2 was performed: Model 1 was fit for each sample. Using the fitted probit regression, C5, C25, C50, C75, and C95 were estimated with an inverse prediction for each sample, and the differences in the intercepts, slopes, and estimated C*X* levels between two samples were recorded. By bootstrap sampling 10,000 times using the R package “boot” (version 1.3-28.1), distributions of the differences in the intercepts, slopes of the probit regressions, and the estimated C*X* levels were obtained. The point estimates and two-sided 95% CIs for differences in intercepts, slopes and C*X* levels were the means and the bias-corrected and accelerated (BCa) (9) intervals from the bootstrap distributions for the corresponding statistics, respectively. Differences were considered statistically different from zero when their 95% CI did not include 0. In addition to the bootstrap approach with the non-centered Model 1 described above, the bootstrap approach with the centered model was also examined, with the only difference being that Model 2 was substituted for Model 1.

Point estimates of differences derived from the interaction term approach (model outputs) and the bootstrap approach (means of the bootstrap distributions of the differences) were recorded for evaluation purpose.

### 2.4. Evaluating the Performance of the Two Approaches Using Simulations

To evaluate the type □ error rate for detecting overall differences between sample types using the interaction term and bootstrap approaches, 1000 simulations under Scenario 1 were performed where data were generated using the same intercepts and slopes (no true difference). Similarly, to evaluate the power of the two approaches under a plausible set of alternatives, 1000 simulations were performed for each of the other four scenarios (Scenarios 2-5) where data were simulated using combinations of different intercepts and/or slopes. In each simulation, VFs *X*_*ij*_ and outcomes *Y*_*ij*_ for a new pair of surrogate and patient samples were generated. Next, analyses using the interaction term approach and the bootstrap approach as described in 2.3.1 and 2.3.2 were performed. This process was repeated 1000 times. Two-sided 95% Wald CIs were computed for differences in intercepts and slopes for the interaction term approach. Two-sided 95% BCa CIs were computed for differences in intercepts, slopes, along with differences in C*X* levels for the bootstrap approach.

Differences in the intercepts and slopes can be obtained in both approaches, allowing for using hypothesis-testing based on these differences to compare performance between approaches. The associated hypotheses for comparing the overall differences between probit regressions of the two sample types are

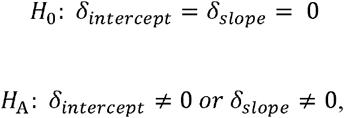

where *δ*_*intercept*_ and *δ*_*slope*_ denote the differences in intercepts and slopes, respectively. The type □ error rate and power were determined as the percentage of 1000 simulations in which the 95% CI for the differences in intercepts or slopes did not include 0.

### 2.5. Evaluating the Performance of Detecting Overall Differences Between Sample Types Based on CX Levels Using Simulations

In addition to quantifying differences in intercepts and slopes, Godsey et al. (7) recommend assessing differences at several C*X* levels (C5, C25, C50, C75, C95) to evaluate differences between sample types. To consider whether differences in C*X* levels (denoted as *δ*_*cx*_) can be utilized to determine overall differences between sample types. Two hypothesis-testing frameworks were investigated. First, overall differences between sample types were accessed by testing the null hypothesis that all differences in the five C*X* levels were 0, that is:

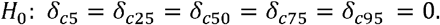

The alternative hypothesis is that at least one of these differences is not 0.

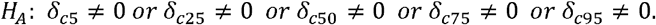

Next, since the C95 level is most commonly used to represent assay performance, the possibility that overall differences between sample types may be determined by evaluating C95 alone was also explored. In this case, the null hypothesis is *H*_0_: *δ*_*c*95_ *=* 0 and the alternative hypothesis is *H*_*A*_: *δ*_*c*95_ ≠ 0. In both setups, *δ*_*cx*_ is determined to be 0 if its two-sided 95% BCa CIs include 0. Type □ error rate and power were computed for each criterion using the same method as described in 2.4.

## 3. Results

### 3.1. A Simulated Example Dataset: Mimicking an NGS-based CSFC Study for Five Scenarios

To evaluate the statistical analysis approaches for a CSFC study for an NGS-based qualitative reporting assay, datasets of six dilution levels, each with 40 replicates, were simulated for a pair of surrogate and patient samples under five scenarios (Table 1). Figure 1 A through E shows the VF distributions across levels for an example dataset for each simulated scenario. Within each level, VFs of the 40 replicates from a sample appear to be normally distributed; across the six levels, as VFs increase, hit rates also increase. Table S1 lists the hit rates and summary statistics of the VFs by level for each sample in the example dataset. To further explore the relationship between the binary outcomes and VFs, a probit regression was fit between them for each sample as depicted in Figure 1 F through J. In Scenario 1, VFs of the two samples were generated based on probit models with the same intercepts and slopes and hence the fitted probit regressions are close to each other. In Scenario 2, VFs of the two samples were simulated based on probit models with the same slopes, but different intercepts. The fitted probit regression of Sample B2, whose VFs were simulated with a smaller intercept, has a parallel shift to the right of A2. In both Scenario 3 and 4, the two samples were simulated using probit models of different intercepts and slopes: A had a larger intercept but a smaller slope, whereas B had a smaller intercept but a larger slope. The difference between Scenario 3 and 4 is that the slope of B3 is so much larger than that of A3 such that the probit regression of B3 surpasses that of A3 near the midpoint, whereas the probit regression of B4 approaches that of A4 but does not surpass A4. Scenario 5 was simulated based on probit models with the same intercepts but different slopes. B5 had a smaller slope than that of A5. Hence, the fitted regression of B5 is close to that of A5 in the low range of VFs but diverges as VF increases.

**Figure 1.**
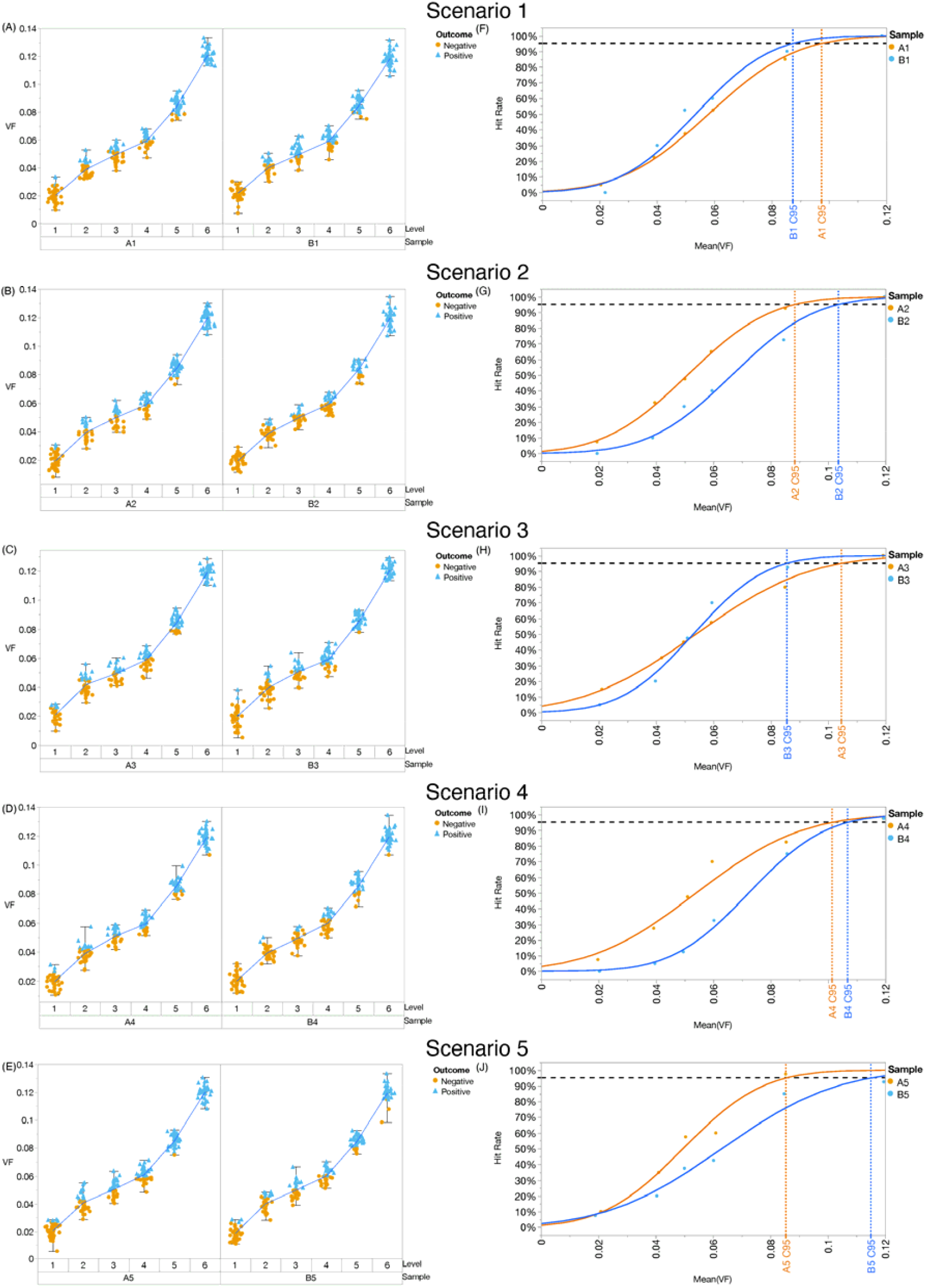
Variant frequencies by level per sample type (left panel) and fitted probit regressions within each sample (right panel). Plots A to E present variant frequencies of a surrogate (A) and patient sample pair (B) across the six dilution levels for the five simulation scenarios. Each of the observations is colored by whether the targeted variant is test positive or negative. Plots F to J show the fitted probit regressions with the non-centered model between the binary outcomes and means of variant frequencies per level across six levels for a pair of surrogate (A, orange line) and patient sample (B, blue line) for the five simulation scenarios.

### 3.2. A Simulated Example Dataset: Fitting a Probit Regression Within Each Sample

Table 2 describes the fitted probit regression, the estimated hit rates derived from forward prediction, and the estimated C*X* levels derived from inverse prediction for each sample in the example dataset. The estimated intercept 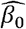 and slope 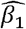 are largely consistent with the parameters used in simulation (Table 1). Because of the high sensitivity inherent in the NGS technology, VFs below C95 are typically small, posing challenges in getting a concrete understanding of practical impacts of differences in C*X* levels. In addition to leveraging the estimated C*X* levels recommended in Godsey et al. (7), another way to compare a pair of surrogate and patient samples is to use the predicted hit rate at a given VF. In Scenario 1 where Surrogate Sample A1 and Patient Sample B1 were simulated using models with the same *β*_0_ and *β*_1_, B1 has a slightly smaller C95 than A1 and thus when the VF is equal to A1’s C95, the estimated hit rate of B1 is 98.35%, slightly higher than 95%; when the VF is equal to B1’s C95, the estimated hit rate of A1 is 88.99%, slightly lower than 95%. However, in a scenario where the surrogate and patient sample were simulated using models with different *β*_0_ and/or *β*_1_, the estimated hit rate of one sample at the other sample’s estimated C95 can be substantially different than 95%. For instance, in Scenario 5, Patient Sample B5 has larger C95 than Surrogate Sample A5. When the VF is equal to A1’s C95, the estimated hit rate of B1 is 76.06%, much lower than 95%.

**Table 2.**
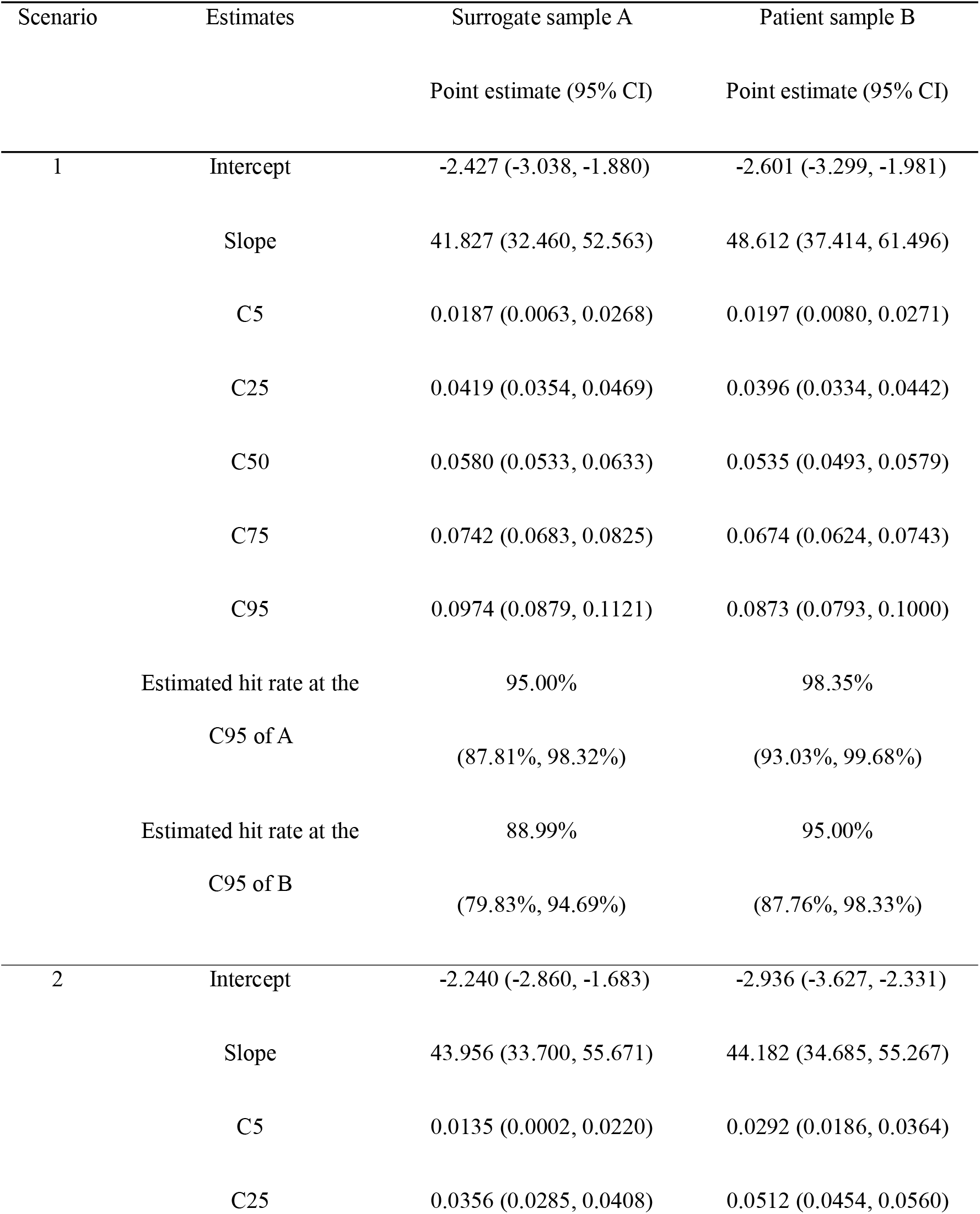
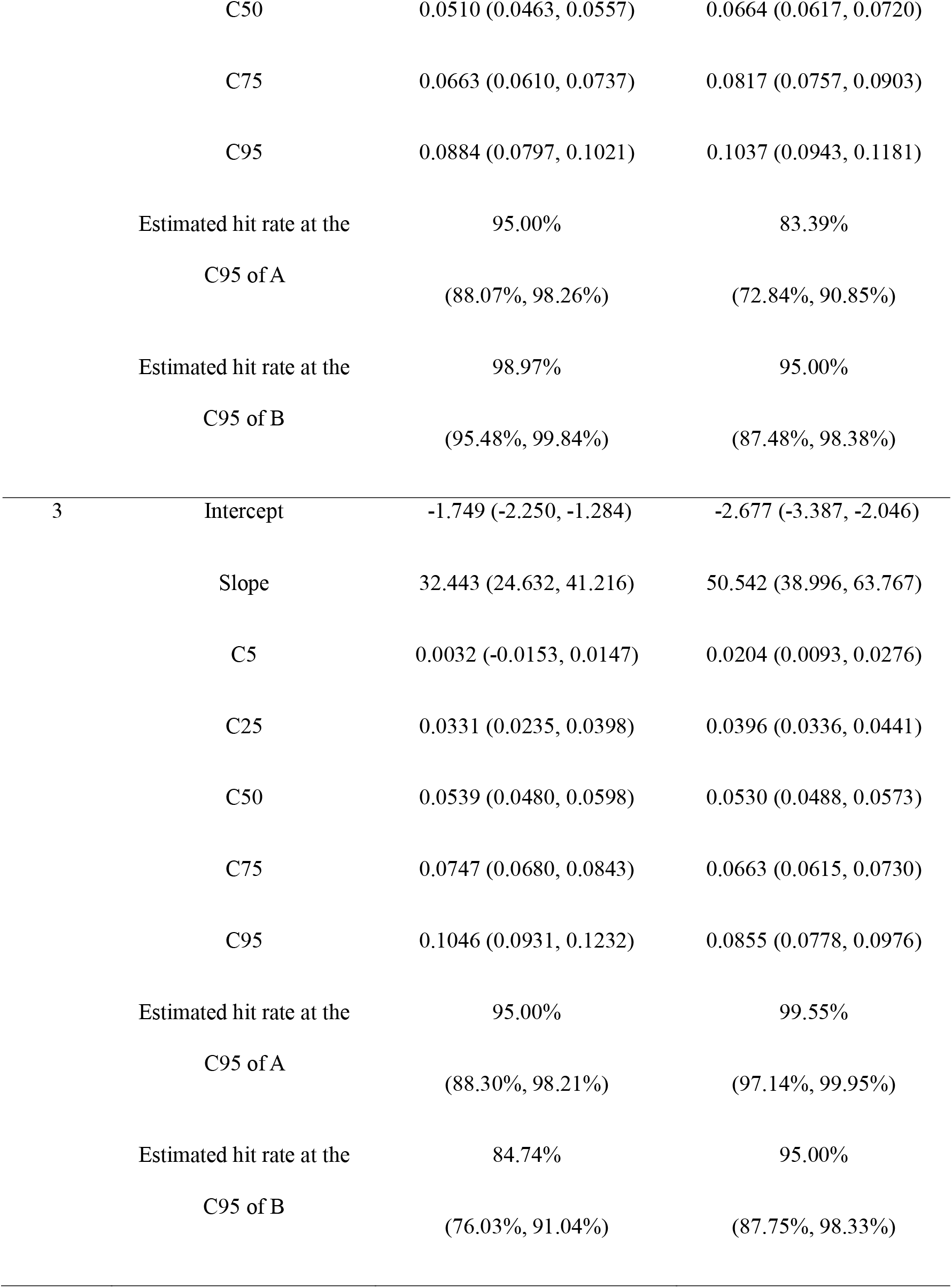
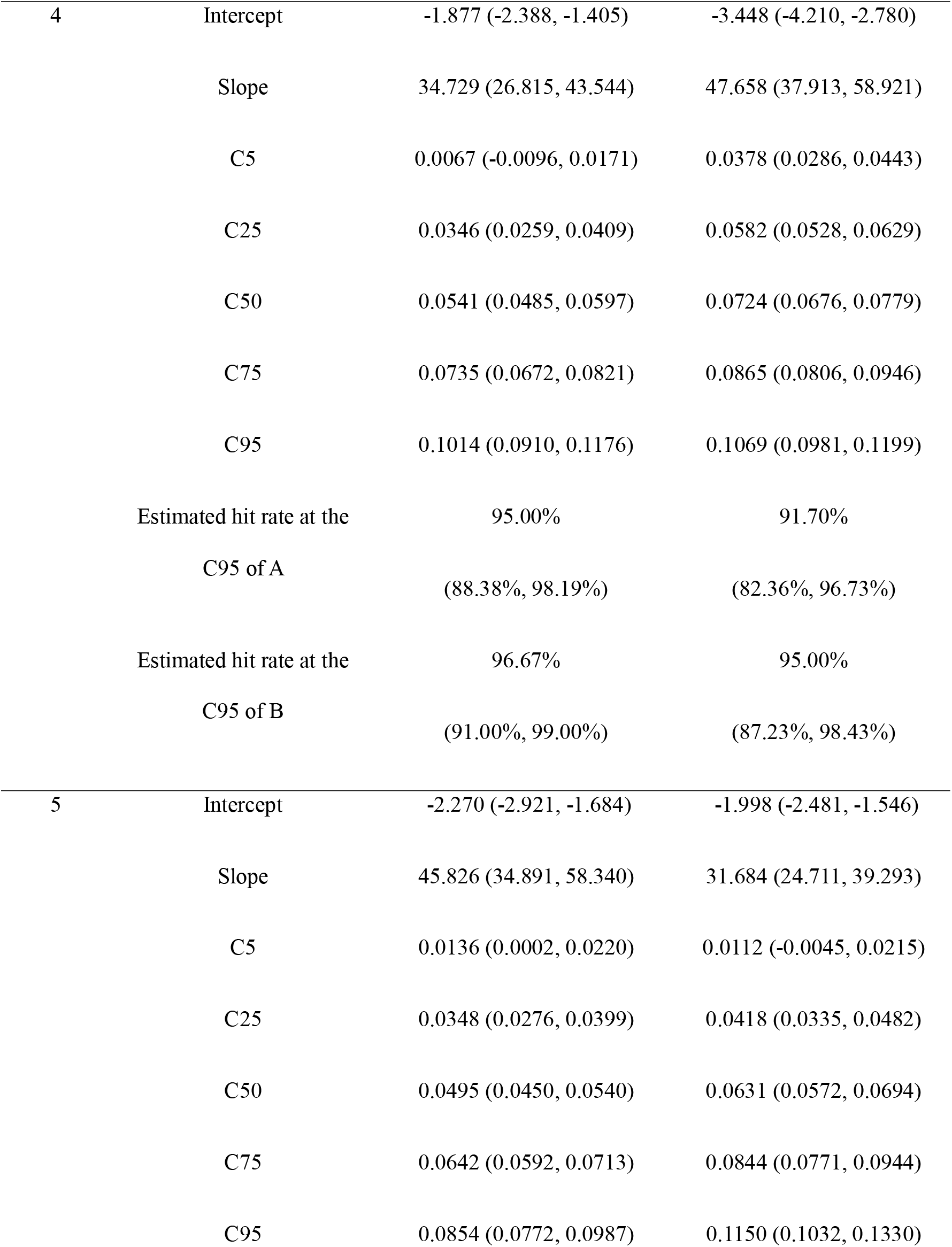
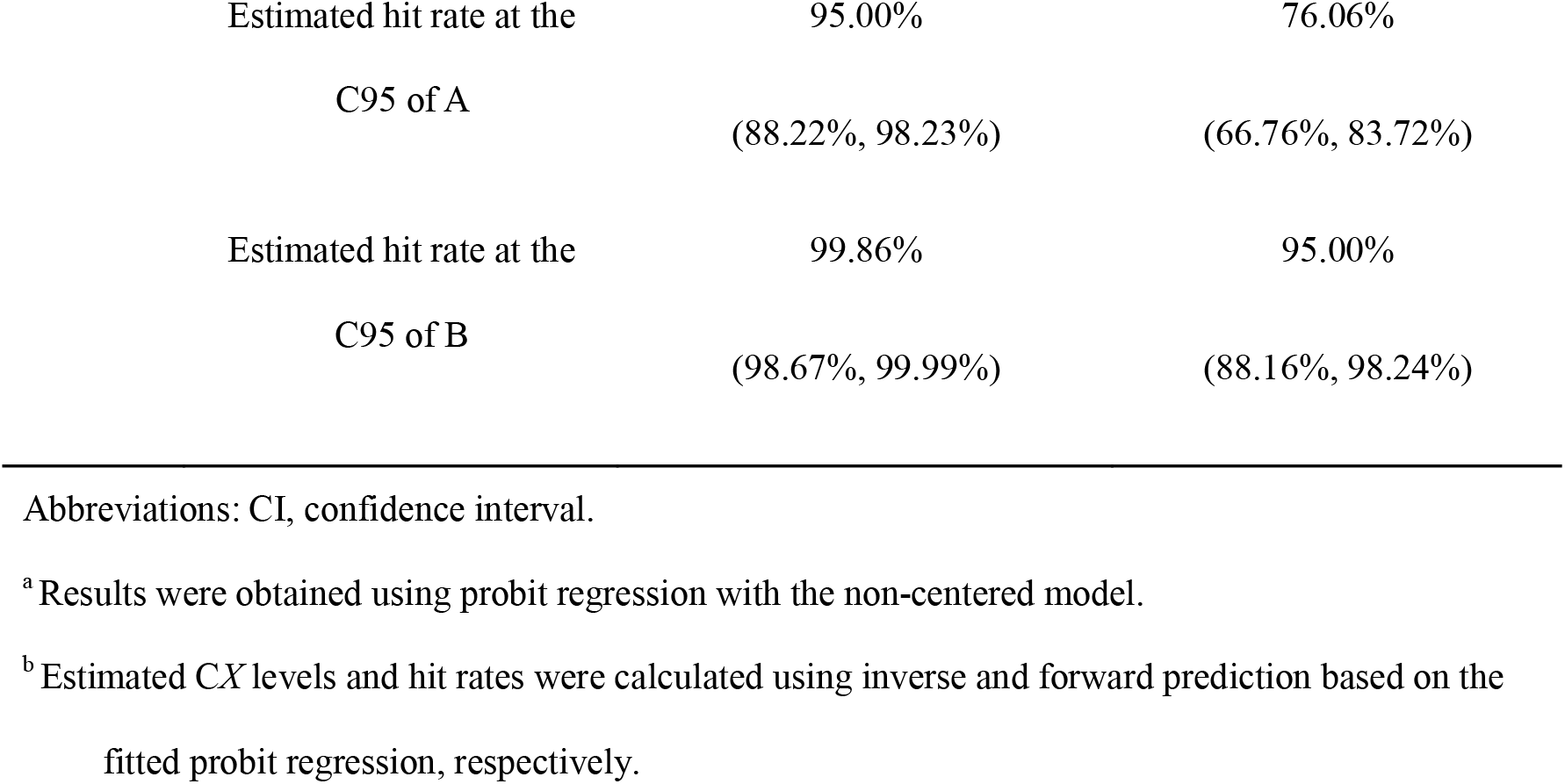
Probit regression^a^ estimates for individual samples and derived predictions^b^.

| Scenario | Estimates | Surrogate sample A | Patient sample B |
| --- | --- | --- | --- |
|  |  | Point estimate (95% CI) | Point estimate (95% CI) |
| 1 | Intercept | -2.427 (-3.038, -1.880) | -2.601 (-3.299, -1.981) |
|  | Slope | 41.827 (32.460, 52.563) | 48.612 (37.414, 61.496) |
|  | C5 | 0.0187 (0.0063, 0.0268) | 0.0197 (0.0080, 0.0271) |
|  | C25 | 0.0419 (0.0354, 0.0469) | 0.0396 (0.0334, 0.0442) |
|  | C50 | 0.0580 (0.0533, 0.0633) | 0.0535 (0.0493, 0.0579) |
|  | C75 | 0.0742 (0.0683, 0.0825) | 0.0674 (0.0624, 0.0743) |
|  | C95 | 0.0974 (0.0879, 0.1121) | 0.0873 (0.0793, 0.1000) |
|  | Estimated hit rate at the | 95.00% | 98.35% |
|  | C95 of A | (87.81%, 98.32%) | (93.03%, 99.68%) |
|  | Estimated hit rate at the | 88.99% | 95.00% |
|  | C95 of B | (79.83%, 94.69%) | (87.76%, 98.33%) |
| 2 | Intercept | -2.240 (-2.860, -1.683) | -2.936 (-3.627, -2.331) |
|  | Slope | 43.956 (33.700, 55.671) | 44.182 (34.685, 55.267) |
|  | C5 | 0.0135 (0.0002, 0.0220) | 0.0292 (0.0186, 0.0364) |
|  | C25 | 0.0356 (0.0285, 0.0408) | 0.0512 (0.0454, 0.0560) |
|  | C50 | 0.0510 (0.0463, 0.0557) | 0.0664 (0.0617, 0.0720) |
|  | C75 | 0.0663 (0.0610, 0.0737) | 0.0817 (0.0757, 0.0903) |
|  | C95 | 0.0884 (0.0797, 0.1021) | 0.1037 (0.0943, 0.1181) |
|  | Estimated hit rate at the | 95.00% | 83.39% |
|  | C95 of A | (88.07%, 98.26%) | (72.84%, 90.85%) |
|  | Estimated hit rate at the | 98.97% | 95.00% |
|  | C95 of B | (95.48%, 99.84%) | (87.48%, 98.38%) |
| 3 | Intercept | -1.749 (-2.250, -1.284) | -2.677 (-3.387, -2.046) |
|  | Slope | 32.443 (24.632, 41.216) | 50.542 (38.996, 63.767) |
|  | C5 | 0.0032 (-0.0153, 0.0147) | 0.0204 (0.0093, 0.0276) |
|  | C25 | 0.0331 (0.0235, 0.0398) | 0.0396 (0.0336, 0.0441) |
|  | C50 | 0.0539 (0.0480, 0.0598) | 0.0530 (0.0488, 0.0573) |
|  | C75 | 0.0747 (0.0680, 0.0843) | 0.0663 (0.0615, 0.0730) |
|  | C95 | 0.1046 (0.0931, 0.1232) | 0.0855 (0.0778, 0.0976) |
|  | Estimated hit rate at the | 95.00% | 99.55% |
|  | C95 of A | (88.30%, 98.21%) | (97.14%, 99.95%) |
|  | Estimated hit rate at the | 84.74% | 95.00% |
|  | C95 of B | (76.03%, 91.04%) | (87.75%, 98.33%) |
| 4 | Intercept | -1.877 (-2.388, -1.405) | -3.448 (-4.210, -2.780) |
|  | Slope | 34.729 (26.815, 43.544) | 47.658 (37.913, 58.921) |
|  | C5 | 0.0067 (-0.0096, 0.0171) | 0.0378 (0.0286, 0.0443) |
|  | C25 | 0.0346 (0.0259, 0.0409) | 0.0582 (0.0528, 0.0629) |
|  | C50 | 0.0541 (0.0485, 0.0597) | 0.0724 (0.0676, 0.0779) |
|  | C75 | 0.0735 (0.0672, 0.0821) | 0.0865 (0.0806, 0.0946) |
|  | C95 | 0.1014 (0.0910, 0.1176) | 0.1069 (0.0981, 0.1199) |
|  | Estimated hit rate at the<br>C95 of A | 95.00%<br>(88.38%, 98.19%) | 91.70%<br>(82.36%, 96.73%) |
|  | Estimated hit rate at the<br>C95 of B | 96.67%<br>(91.00%, 99.00%) | 95.00%<br>(87.23%, 98.43%) |
| 5 | Intercept | -2.270 (-2.921, -1.684) | -1.998 (-2.481, -1.546) |
|  | Slope | 45.826 (34.891, 58.340) | 31.684 (24.711, 39.293) |
|  | C5 | 0.0136 (0.0002, 0.0220) | 0.0112 (-0.0045, 0.0215) |
|  | C25 | 0.0348 (0.0276, 0.0399) | 0.0418 (0.0335, 0.0482) |
|  | C50 | 0.0495 (0.0450, 0.0540) | 0.0631 (0.0572, 0.0694) |
|  | C75 | 0.0642 (0.0592, 0.0713) | 0.0844 (0.0771, 0.0944) |
|  | C95 | 0.0854 (0.0772, 0.0987) | 0.1150 (0.1032, 0.1330) |

|  |  |  |
| --- | --- | --- |
| Estimated hit rate at the | 95.00% | 76.06% |
| C95 of A | (88.22%, 98.23%) | (66.76%, 83.72%) |
| Estimated hit rate at the | 99.86% | 95.00% |
| C95 of B | (98.67%, 99.99%) | (88.16%, 98.24%) |

Abbreviations: CI, confidence interval.
<sup>a</sup> Results were obtained using probit regression with the non-centered model.
<sup>b</sup> Estimated CX levels and hit rates were calculated using inverse and forward prediction based on the fitted probit regression, respectively.

### 3.3. A Simulated Example Dataset: Generating CIs for Differences Between Sample Types Using the Interaction Term and Bootstrap Approaches

As suggested by Godsey et al. (7), CIs for differences in intercepts and slopes of probit regressions, as well as differences in estimated C*X* levels can be used to evaluate the differences between sample types. The interaction term and bootstrap approaches were evaluated to generate CIs for the differences, and both approaches were assessed with the non-centered and centered model, as described in 2.3.1 and 2.3.2. Because of the nature of a probit model, the slope estimate is always the same whether fitted with the non-centered or centered model. The difference between the non-centered and centered model is that the non-centered model tests differences in intercepts when VF equals zero, whereas the centered model tests differences in intercepts when VF is at the mean VF across replicates spanning the six levels of the two samples.

Results from the interaction term approach are shown in the left section of Table 3. In Scenario 1, no statistically significant differences were detected in the intercepts or slopes. This is expected because Surrogate Sample A and Patient Sample B were generated from the same probit model. In Scenario 2, the centered model detected difference in the intercepts, whereas the non-centered model did not detect any difference in the intercepts or slopes. In Scenarios 3 to 5, at least one of the intercepts and slopes was found to be statistically different (two-sided 95% CIs did not include 0). In Scenario 3, the non-centered model detected statistical difference for the intercept while the centered model did not. This is expected since the two samples in Scenario 3 intersect in the middle, leading to minimal differences in intercepts around that point, making it challenging for the centered model to detect these differences. Nonetheless, the difference in slopes in scenario 3 was detected using either model. In Scenario 4, both models detected the differences in the intercept, but not the slope. This is reasonable given that the slope difference in Scenario 4 is smaller than that in Scenario 3. In Scenario 5, the centered model detected statistical difference for the intercept while the non-centered model did not. This is consistent with the observation that the two samples have diverged substantially since they overlapped at the origin, providing the centered model with greater statistical power for testing differences in intercepts. The difference in slopes is of borderline significance.

**Table 3.**
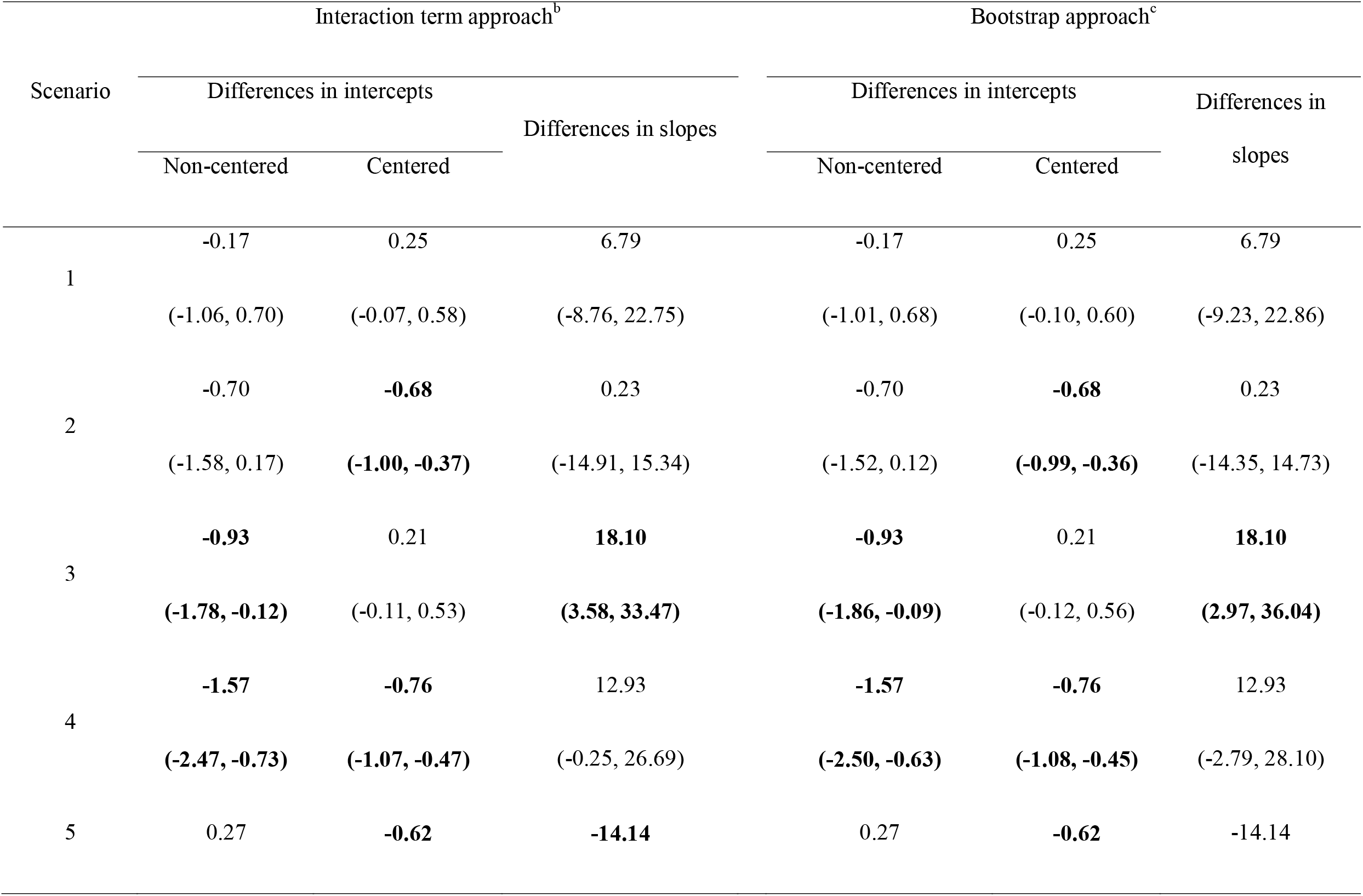
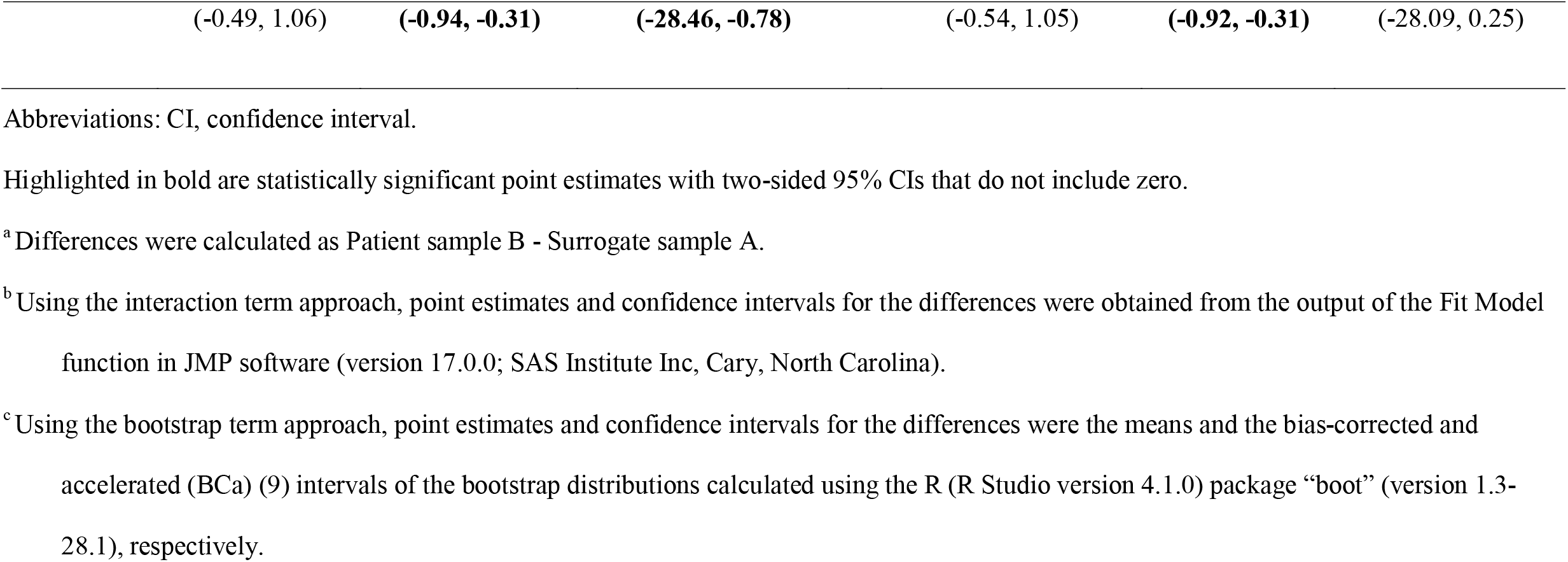
Comparison of differences^a^ between probit regression from the example dataset using the interaction term and bootstrap approaches.

| Scenario | Interaction term approach<sup>b</sup> |  |  | Bootstrap approach<sup>c</sup> |  |  |
| --- | --- | --- | --- | --- | --- | --- |
|  | Differences in intercepts |  | Differences in slopes | Differences in intercepts |  | Differences in slopes |
|  | Non-centered | Centered |  | Non-centered | Centered |  |
| 1 | -0.17 | 0.25 | 6.79 | -0.17 | 0.25 | 6.79 |
|  | (-1.06, 0.70) | (-0.07, 0.58) | (-8.76, 22.75) | (-1.01, 0.68) | (-0.10, 0.60) | (-9.23, 22.86) |
| 2 | -0.70 | **-0.68** | 0.23 | -0.70 | **-0.68** | 0.23 |
|  | (-1.58, 0.17) | **(-1.00, -0.37)** | (-14.91, 15.34) | (-1.52, 0.12) | **(-0.99, -0.36)** | (-14.35, 14.73) |
| 3 | **-0.93** | 0.21 | **18.10** | **-0.93** | 0.21 | **18.10** |
|  | **(-1.78, -0.12)** | (-0.11, 0.53) | **(3.58, 33.47)** | **(-1.86, -0.09)** | (-0.12, 0.56) | **(2.97, 36.04)** |
| 4 | **-1.57** | **-0.76** | 12.93 | **-1.57** | **-0.76** | 12.93 |
|  | **(-2.47, -0.73)** | **(-1.07, -0.47)** | (-0.25, 26.69) | **(-2.50, -0.63)** | **(-1.08, -0.45)** | (-2.79, 28.10) |
| 5 | 0.27 | **-0.62** | **-14.14** | 0.27 | **-0.62** | -14.14 |

|  |  |  |  |  |  |
| --- | --- | --- | --- | --- | --- |
| (-0.49, 1.06) | **(-0.94, -0.31)** | **(-28.46, -0.78)** | (-0.54, 1.05) | **(-0.92, -0.31)** | (-28.09, 0.25) |

Abbreviations: CI, confidence interval.
Highlighted in bold are statistically significant point estimates with two-sided 95% CIs that do not include zero.
<sup>a</sup> Differences were calculated as Patient sample B - Surrogate sample A.
<sup>b</sup> Using the interaction term approach, point estimates and confidence intervals for the differences were obtained from the output of the Fit Model function in JMP software (version 17.0.0; SAS Institute Inc, Cary, North Carolina).
<sup>c</sup> Using the bootstrap term approach, point estimates and confidence intervals for the differences were the means and the bias-corrected and accelerated (BCa) (9) intervals of the bootstrap distributions calculated using the R (R Studio version 4.1.0) package “boot” (version 1.3-28.1), respectively.

Another way of constructing CIs for differences in intercepts, slopes of probit regressions, and estimated C*X* levels is from generated bootstrap distributions, as described in 2.3.2. Similar to the interaction term approach, the bootstrap approach was implemented with both the centered and non-centered models. As shown in the right section of Table 3, the bootstrap approach yielded results closely aligning with those from the interaction term approach, consistent across both the non-centered and centered models. As expected, the point estimates of the differences in the intercepts and slopes are identical between the two approaches. The slight difference in the CIs can be attributed to the two different approaches, different methods used to calculate CIs, or both.

The bootstrap approach allows us to directly derive CIs not just for the differences in intercepts and slopes of probit regressions, but also for the estimated C*X* levels, with results shown in Table 4.

**Table 4.**
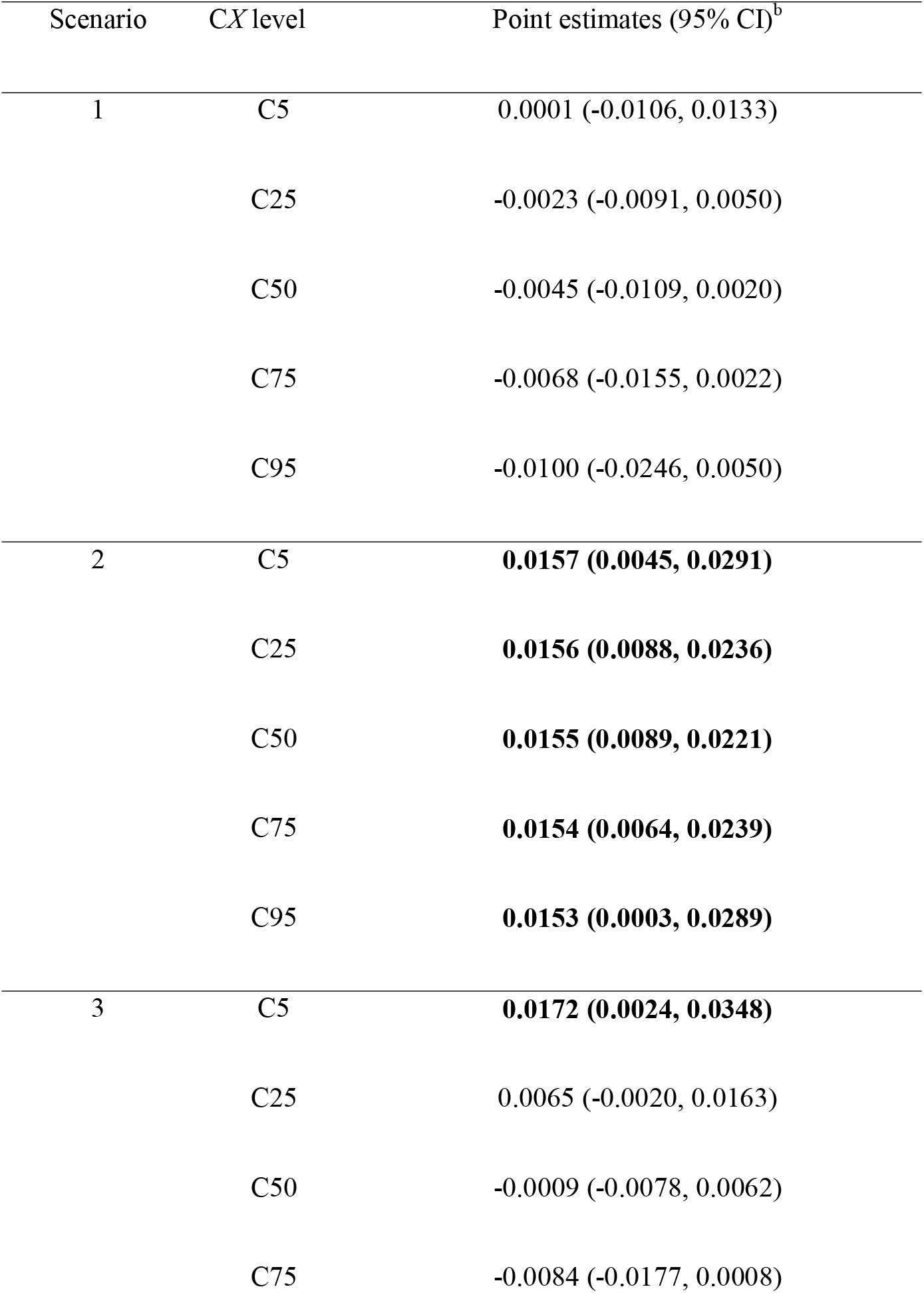
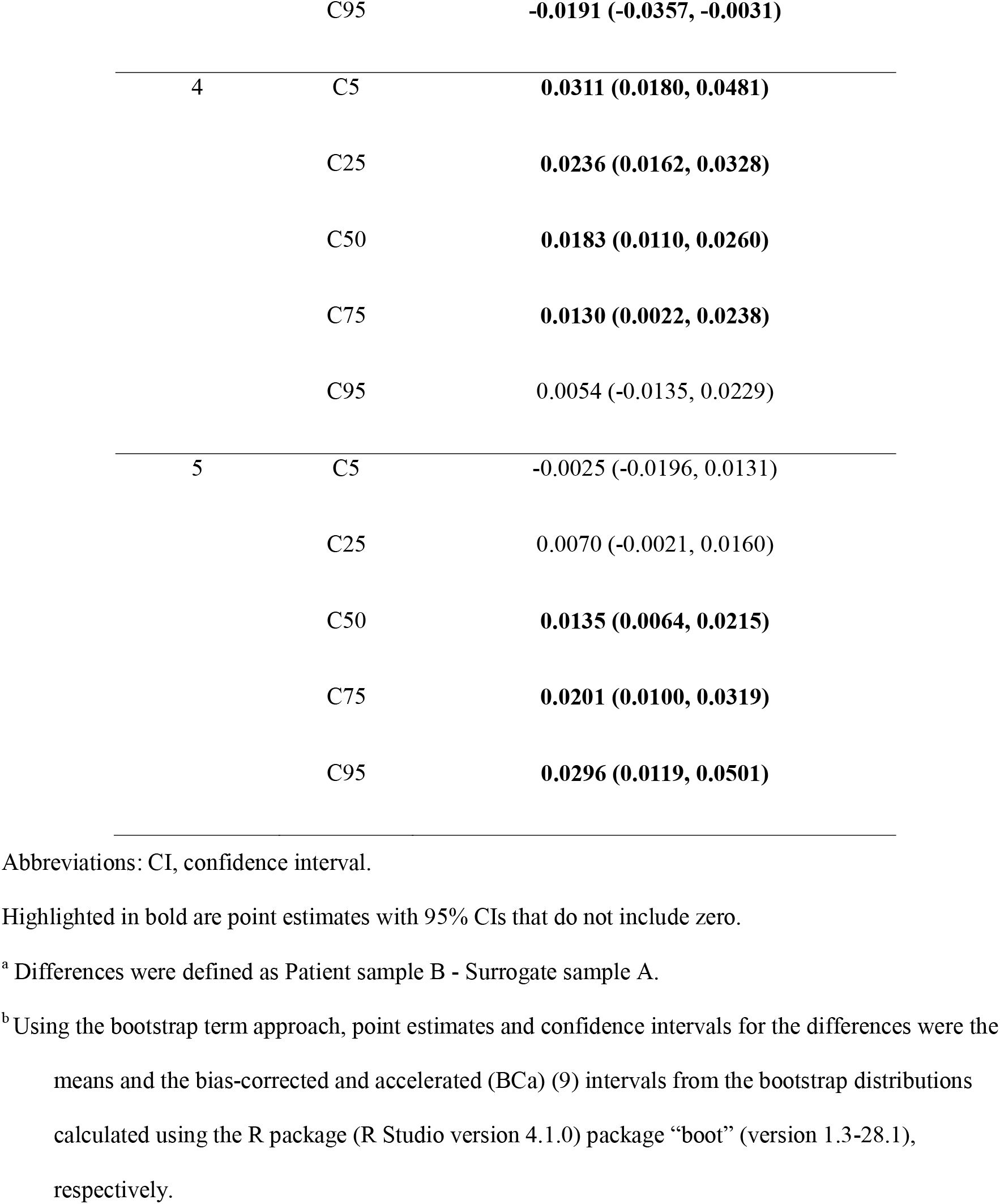
Differences^a^ in estimated C*X* levels obtained by applying the bootstrap approach to the example data.

Using either the non-centered or centered models in each bootstrap run does not alter estimated C*X* levels derived from inverse predictions. Consequently, it does not affect point estimates or CIs for differences in estimated C*X* levels. In Scenario 1, as expected, all the estimated C*X* levels had 95% CIs including zero. For Scenario 2, where the probit regressions were simulated using the same slopes but different intercepts, it can be easily derived that the differences in C*X* levels should be constant across C*X* levels. This consistency is reflected in the minimal variation observed in the estimated differences in C5, C25, C50, C75 and C95 of the two samples in the example dataset. In Scenarios 3 where the probit regressions intersect in the middle and deviate most from each other in both ends, it is consistent to observe that the 95% CIs for C5 and C95 do not include zero. In Scenario 4, as the VF increases, the probit regressions converge, and it is consistent to observe that the 95% CIs for C95 include zero. Similarly in scenario 5 where the probit regressions diverge as the VF increases, it is consistent to see that C50, C75 and C95 have 95% CIs intervals that do not include zero. In general, the statistical significance of 95% CIs for the differences in estimated C*X* levels mirrors closely with where the probit regressions differ most.

### 3.4. 1000 Simulated Datasets: Evaluating Test Performance of the Interaction Term and Bootstrap Approaches

To compare the type □ error rate and power of the interaction term and bootstrap approaches in detecting overall differences in probit regressions, 1000 simulations were performed for the five scenarios using both the non-centered and centered models (Table 5). Across all five scenarios, the bootstrap approach demonstrated similar performance as the interaction term approach for both the non-centered and centered models. Regarding type □ error, the hypotheses for comparing overall differences between probit regressions of the two sample types involve tests for both the intercept and slope. Assuming each test is well-calibrated, the type □ error rate is expected to be bounded at 10% (5% × 2). All observed type □ error rates are consistent with expectations. Under Scenario 1, with the non-centered model, the interaction term model approach had 61 simulations where differences in the intercepts or slopes showed 95% CIs not including 0, indicating a type □ error rate of 6.1%. The bootstrap approach showed a similar type □ error rate of 5.4%. With the centered model, both the interaction term and bootstrap approaches had slightly higher type □ error rates of 9% and 7.8%, respectively. Power was evaluated in the other four scenarios where data was simulated using probit models with different intercepts and/or slopes. When the non-centered model was applied, the power of the interaction term approach was low for Scenario 2 (17.3%), modest for Scenario 5 (60.8%), and good for Scenario 3 (94.5%) and Scenario 4 (98.2%). However, when the centered model was applied, the power was good for all scenarios (88.5% in Scenario 2, 90.4% in Scenario 3, 100% in both Scenario 4 and 5). Using both non-centered and centered models, the bootstrap approach showed comparable power as the interaction term approach.

**Table 5.**
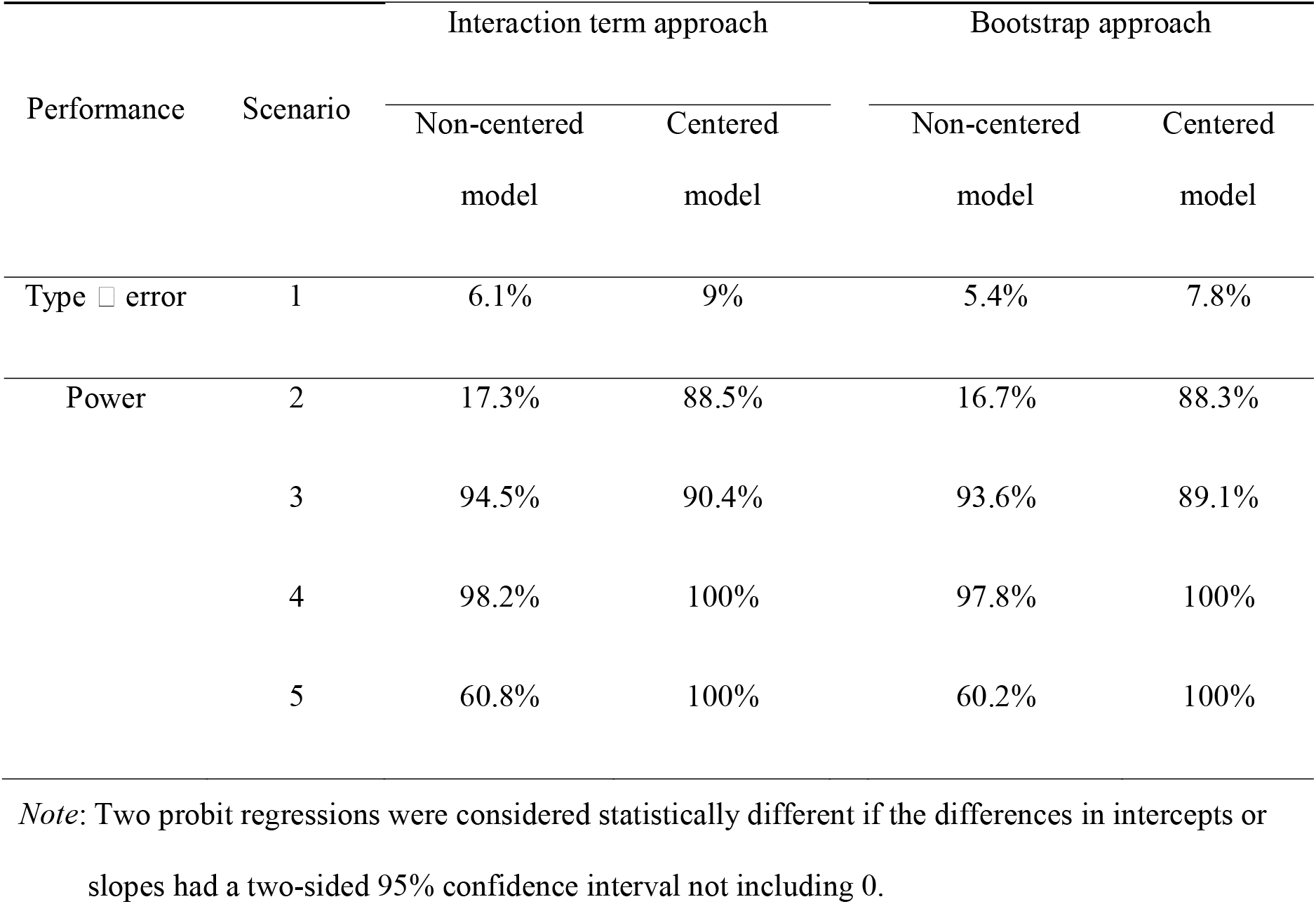
Performance comparison between the interaction term and bootstrap approaches based on probit model parameters from 1000 simulations.

### 3.5. 1000 Simulated Datasets: Evaluating Performance for Detecting Overall Differences Between Sample Types Based on CX Levels

As an alternative to using differences in intercepts or slopes to detect overall differences between sample types, the performance of using differences in C*X* estimates obtained from the bootstrap approach was evaluated with two frameworks. In the first framework, the overall probit regressions between sample types were considered different when differences in at least one of C5, C25, C50, C75, and C95 were not equal to 0, as defined by 95% CIs not including 0. Since this framework involves five tests, a higher type □ error rate is anticipated, with an upper bound of 25% (5% × 5) assuming each test is well-calibrated. As shown in Table 6, the type □ error rate was relatively high at 16.4%. The power is good for the other four scenarios (94.7% in Scenario 2, 96.9% in Scenario 3, and 100% in both Scenario 4 and 5). Since C95 is a key metric for accessing assay performance, the performance to detect overall differences based on C95 alone was also evaluated. This resulted in a lower type □error rate (6%). Power is good for Scenario 5 (99.4%), modest for Scenario 3 (69.5%), but low for Scenario 2 (31.4%) and Scenario 4 (11.0%).

**Table 6.** Comparison of two hypothesis-testing frameworks using C*X* estimates obtained via the bootstrap approach from 1000 simulations.

| Alternative Hypothesis<sup>a</sup> | Type I error rate | Power |  |  |  |
| --- | --- | --- | --- | --- | --- |
|  | Scenario 1 | Scenario 2 | Scenario 3 | Scenario 4 | Scenario 5 |
| At least one of the differences in CX levels not equal to 0<sup>b</sup> | 16.4% | 94.7% | 96.9% | 100% | 100% |
| Differences in C95 not equal to 0<sup>b</sup> | 6% | 31.4% | 69.5% | 11.0% | 99.4% |

<sup>a</sup>Two hypothesis-testing frameworks were investigated to evaluate the performance of detecting overall differences between sample types based on differences in CX levels. First, overall differences between sample types were accessed by testing the null hypothesis that all differences in the five CX levels were 0. Second, the null hypothesis was that the difference in C95 alone was not 0.
<sup>b</sup>Differences in CX levels were determined to be 0 if their two-sided 95% bias-corrected and accelerated (BCa) (9) confidence intervals included 0.

## 4. Discussion

The use of surrogate samples ameliorates the difficulty of lacking patient samples in assay development and validation. When selecting surrogate samples, it is crucial to thoroughly consider sample properties that impact the detection performance of a medical device following CLSI EP39 (4). One aspect of this assessment for qualitative reporting assays may involve a thorough evaluation of the comparability between samples through a CSFC study. In a sufficiently powered CSFC study, the absence of statistically significant differences in the probit regressions suggests that the surrogate sample is comparable to the patient sample in terms of how the detection performance responds to the change in the concentration of the target variant. However, additional testing comparing various other aspects may be necessary to further assess the appropriateness of using surrogate samples to augment patient samples. Conversely, in the presence of a statistically significant difference between samples, investigations are needed to identify the causes. Differences in factors such as operator and reagent lot between surrogate and patient samples, if not held constant, may contribute to the differences. Additionally, statistically significant differences necessitate the assessment of whether such a degree of difference will result in practically meaningful impacts. A benefit-risk analysis that evaluates the proportion of the intended-use population affected by the differences is helpful for assay developers to better understand the impact of these differences.

To establish robust analysis methods to carry out the statistical evaluation of CSFC studies proposed by Godsey et al. (7), we evaluated the interaction term and bootstrap approaches in detecting differences in intercepts and slopes between probit regressions using both the non-centered and centered models across five simulated scenarios. The interaction term approach is a parametric approach that assumes the probability of a binary outcome is determined by a normal latent variable, which is modeled as a linear combination of predictors and a standard normal error term (10). Therefore, this approach may not be suitable for a CSFC study if these underlying assumptions are violated in the data. In contrast, the bootstrap approach does not assume a specific parametric form for the distribution, making it more flexible in such cases. In terms of ease of implementation, the interaction term approach is computationally less burdensome compared to the bootstrap approach. In terms of performance, as revealed consistently across scenarios, the interaction term and bootstrap approaches showed similar results for the simulated data considered, whether using the non-centered or centered model.

Compared to the non-centered model, the centered model is theoretically better suited for detecting hit rate differences in a CSFC study for NGS-based assays. This is because such differences typically become more pronounced near the middle of the probit curves, where comparisons are more clinically meaningful. In terms of performance, it is evident that the centered model was effective in detecting overall differences between probit regression for all scenarios, performing better than the non-centered model in Scenario 2 and 5. In Scenarios 2 and 5, the limited ability of the non-centered model to detect true differences was at least partly attributed to the expected zero hit rate when the VF was zero, resulting in no discernible difference in hit rates. In scenario 3, where probit regressions intersected near the midpoint, the substantial difference in slopes made it easier to detect differences in slopes. As a result, the non-centered and centered models exhibited similar performance driven by the detection of differences in slopes. In scenario 4, the non-centered model was able to detect the small differences in intercepts when the VF was zero, therefore, only a slight decrease in performance was seen with the non-centered model compared to the centered model. Collectively, the centered model is recommended over the non-centered model for CSFC data similar to that simulated in this work.

Upon reviewing the differences in estimated C*X* levels obtained from the bootstrap approach, it was observed that relying solely on C95 to compare sample types often failed to capture the differences, except for Scenario 5. As shown in Figure 1J, the probit regressions diverge as the VF increases in Scenario 5, leading to a large difference in C95 that is more readily detectable. Simultaneously examining the differences in multiple C*X* levels was effective in detecting differences in probit regressions between sample types in all scenarios but greatly elevated the multiple testing burden.

This work compares the use of the interaction term and bootstrap approaches with both the non-centered and centered models to identify an appropriate method for implementing the analysis framework proposed by Godsey et al. (7). Five simulated scenarios, designed to reflect a realistic NGS-based CSFC study, are used to support the evaluation and inform the resulting recommendations. It is worth noting that the VF of replicate j at each dilution level *i* (*X*_*ij*_) was simulated around the same underlying VF for both the surrogate and patient samples. When conducting the experiment, another question to consider is how to ensure the surrogate and patient samples have the same VF for the targeted variant at each dilution level, especially the top-level. Godsey et al. (7) recommends performing quantification for the top-level dilutions using an orthogonal method. However, if the orthogonal method performs differently between sample types, one must consider the possible introduction of bias to the CSFC study. The present study has several limitations. First, probit regression is used throughout this work since CLSI EP17 (8) details the use of probit regression for determining LOD. However, logistic regression, which uses the logistic distribution as the link function, may be used in a CSFC study for qualitative reporting assays instead. Second, due to the lack of publicly available real-word NGS-based CSFC data, our findings are limited to the simulated scenarios used in this study. Future work evaluating the two approaches in other simulation settings will help provide suggestions in a broader scenario. In addition, other ways to statistically evaluate differences in intercepts, slopes, and estimated C*X* levels between samples can be explored, such as construction estimators for the variabilities of these differences using the Delta method.

## 5. Conclusions

Taken together, for data that is similar to our simulated setup, we recommend that the statistical assessment of a CSFC study focus primarily on the differences in intercepts and slopes between probit regressions fitted with the centered model to obtain an assessment of the overall differences, rather than on differences in estimated C*X* levels. While both approaches have strengths and limitations, we recommend using the interaction term approach with the centered model as the primary method due to the ease of computation. The bootstrap approach may offer advantages if C*X* levels are of interest, if pooling surrogate and patient samples is inappropriate, or solely for the purpose of verify findings from the parametric models. Our recommendations are summarized in the simplified guide presented in the supplementary material.

## Supplementary material

### Statistical evaluation recommendations for CSFC studies

#### Part 1: Perform analysis within each sample type

(1) For all replicates at each level and sample type, calculate the mean VF, the number of attempts and hits, and the hit rate (proportion of replicates that test positive).
(2) Fit a centered^a^ probit regression model separately for each sample type using the hits, attempts, and mean VFs calculated in step 1.
(3) Verify model appropriateness by checking diagnostics and goodness of fit metrics^b,c^ .
(4) Using the appropriately fitted probit model for each sample type:
  (a) Obtain the model parameters (intercepts and slopes).
  (b) Estimate the C*X* levels (C5, C25, C50, C75, and C95) through inverse prediction.

#### Part2: Compare parameters between sample types

##### Part2a: Compare intercepts and slopes between sample types

(5) Obtain point estimates of differences in intercepts and slopes by subtraction from step 4a.
(6) Obtain confidence intervals (CIs) for differences in intercepts and slopes:
  (a) If comparing C*X* levels (Part 2b) is not of interest, use the interaction term approach with the centered model^a^ to get CIs for the differences in intercepts and slopes unless the interaction term approach is deemed inappropriate (e.g., surrogate and patient sample data are not poolable^d^, or model diagnostics and goodness of fit tests fail).
  (b) If comparing C*X* levels is of interest or the interaction term approach is deemed inappropriate in step 6a, use the bootstrap approach with the centered model^a^ to get CIs for the differences in intercepts and slopes.

##### Part2b (optional): Compare C*X* levels between sample types

(7) Obtain point estimates of differences in C*X* levels by subtraction from step 4b.
(8) Obtain CIs of differences in estimated C*X* levels using the bootstrap approach with the centered model^a^.

## Declarations

### Ethics approval and consent to participate

Not applicable.

### Consent for publication

Not applicable.

### Data availability

The data supporting the findings of this study were simulated, and the data and code used for simulation and analysis are available from the corresponding author upon reasonable request.

## Acknowledgments

The authors gratefully thank Dawn Erlandson for reviewing the article and providing comments.

## Author contributions

All authors researched the regulatory requirements, concepted and designed this study. LG simulated data and conducted the statistical analysis. All authors interpreted the results. LG drafted the manuscripts. PW and KM substantially revised the manuscript. All authors reviewed and approved the manuscript.

## Abbreviations

BCa: Bias-corrected and accelerated
CDx: Companion diagnostic
CI: Confidence interval
CLSI: Clinical and Laboratory Standards Institute
CSFC: Contrived sample functional characterization
C*X*: Estimated concentrations of the measurand at which an assay declares a sample positive X% of the time
FDA: Food and Drug Administration
LOB: Limit of blank
LOD: Limit of detection
NGS: Next-generation sequencing
PCR: Polymerase chain reaction
TSO: Trusight™ Oncology
VF: Variant allele frequency

---

**Table S1.** Summary statistics for variant frequencies by level for each sample in the simulated example data.

| Scenario | Sample | Level | Hits | Hit<br>Rate<sup>a</sup> | Mean | Median | Min | Max | Range | Std | CV |
| --- | --- | --- | --- | --- | --- | --- | --- | --- | --- | --- | --- |
| 1 | A1 | 1 | 2 | 5% | 0.020 | 0.020 | 0.010 | 0.033 | 0.024 | 0.005 | 26 |
|  |  | 2 | 9 | 23% | 0.039 | 0.038 | 0.032 | 0.053 | 0.021 | 0.005 | 12 |
|  |  | 3 | 15 | 38% | 0.050 | 0.050 | 0.038 | 0.060 | 0.022 | 0.005 | 11 |
|  |  | 4 | 21 | 53% | 0.060 | 0.059 | 0.047 | 0.068 | 0.021 | 0.004 | 7 |
|  |  | 5 | 34 | 85% | 0.085 | 0.085 | 0.074 | 0.095 | 0.021 | 0.005 | 6 |
|  |  | 6 | 40 | 100% | 0.121 | 0.120 | 0.113 | 0.134 | 0.020 | 0.005 | 4 |
|  | B1 | 1 | 0 | 0% | 0.022 | 0.022 | 0.007 | 0.030 | 0.023 | 0.004 | 20 |
|  |  | 2 | 12 | 30% | 0.040 | 0.040 | 0.030 | 0.051 | 0.020 | 0.005 | 12 |
|  |  | 3 | 21 | 53% | 0.050 | 0.048 | 0.038 | 0.063 | 0.025 | 0.006 | 12 |
|  |  | 4 | 24 | 60% | 0.059 | 0.060 | 0.046 | 0.070 | 0.024 | 0.005 | 9 |
|  |  | 5 | 36 | 90% | 0.085 | 0.084 | 0.075 | 0.096 | 0.021 | 0.005 | 6 |
|  |  | 6 | 40 | 100% | 0.118 | 0.118 | 0.106 | 0.132 | 0.026 | 0.005 | 5 |
| 2 | A2 | 1 | 3 | 8% | 0.019 | 0.018 | 0.008 | 0.031 | 0.023 | 0.005 | 28 |
|  |  | 2 | 13 | 33% | 0.040 | 0.040 | 0.028 | 0.050 | 0.022 | 0.005 | 13 |
|  |  | 3 | 19 | 48% | 0.050 | 0.051 | 0.040 | 0.062 | 0.022 | 0.006 | 11 |
|  |  | 4 | 26 | 65% | 0.059 | 0.060 | 0.049 | 0.067 | 0.018 | 0.005 | 8 |
|  |  | 5 | 37 | 93% | 0.085 | 0.086 | 0.073 | 0.094 | 0.021 | 0.004 | 5 |
|  |  | 6 | 40 | 100% | 0.120 | 0.121 | 0.108 | 0.130 | 0.022 | 0.005 | 4 |
|  | B2 | 1 | 0 | 0% | 0.019 | 0.019 | 0.012 | 0.029 | 0.017 | 0.004 | 22 |
|  |  | 2 | 4 | 10% | 0.039 | 0.038 | 0.029 | 0.049 | 0.020 | 0.004 | 10 |
|  |  | 3 | 12 | 30% | 0.050 | 0.050 | 0.041 | 0.059 | 0.018 | 0.004 | 8 |
|  |  | 4 | 16 | 40% | 0.060 | 0.059 | 0.051 | 0.068 | 0.017 | 0.004 | 7 |
|  |  | 5 | 29 | 73% | 0.084 | 0.085 | 0.073 | 0.091 | 0.017 | 0.005 | 5 |
|  |  | 6 | 40 | 100% | 0.120 | 0.120 | 0.107 | 0.135 | 0.028 | 0.006 | 5 |
| 3 | A3 | 1 | 6 | 15% | 0.021 | 0.022 | 0.010 | 0.029 | 0.019 | 0.004 | 21 |
|  |  | 2 | 14 | 35% | 0.042 | 0.043 | 0.030 | 0.056 | 0.027 | 0.006 | 14 |
|  |  | 3 | 18 | 45% | 0.050 | 0.050 | 0.041 | 0.060 | 0.019 | 0.005 | 10 |
|  |  | 4 | 23 | 58% | 0.059 | 0.061 | 0.046 | 0.069 | 0.023 | 0.005 | 9 |
|  |  | 5 | 32 | 80% | 0.085 | 0.085 | 0.077 | 0.095 | 0.018 | 0.005 | 5 |
|  |  | 6 | 40 | 100% | 0.119 | 0.120 | 0.111 | 0.129 | 0.018 | 0.004 | 3 |
|  | B3 | 1 | 2 | 5% | 0.020 | 0.021 | 0.005 | 0.038 | 0.033 | 0.007 | 35 |
|  |  | 2 | 8 | 20% | 0.040 | 0.039 | 0.026 | 0.055 | 0.029 | 0.006 | 16 |
|  |  | 3 | 19 | 48% | 0.051 | 0.051 | 0.040 | 0.064 | 0.024 | 0.005 | 10 |
|  |  | 4 | 28 | 70% | 0.059 | 0.059 | 0.047 | 0.071 | 0.024 | 0.006 | 9 |
|  |  | 5 | 37 | 93% | 0.086 | 0.086 | 0.078 | 0.093 | 0.015 | 0.005 | 5 |
|  |  | 6 | 40 | 100% | 0.120 | 0.120 | 0.114 | 0.129 | 0.016 | 0.004 | 3 |
| 4 | A4 | 1 | 3 | 8% | 0.020 | 0.020 | 0.010 | 0.031 | 0.021 | 0.005 | 24 |
|  |  | 2 | 11 | 28% | 0.039 | 0.040 | 0.027 | 0.057 | 0.030 | 0.005 | 14 |
|  |  | 3 | 19 | 48% | 0.051 | 0.051 | 0.042 | 0.059 | 0.017 | 0.005 | 9 |
|  |  | 4 | 28 | 70% | 0.060 | 0.059 | 0.051 | 0.069 | 0.018 | 0.005 | 8 |
|  |  | 5 | 33 | 83% | 0.085 | 0.086 | 0.076 | 0.099 | 0.023 | 0.004 | 5 |
|  |  | 6 | 39 | 98% | 0.119 | 0.120 | 0.107 | 0.130 | 0.023 | 0.005 | 4 |
|  | B4 | 1 | 0 | 0% | 0.020 | 0.020 | 0.012 | 0.032 | 0.020 | 0.005 | 26 |
|  |  | 2 | 2 | 5% | 0.040 | 0.040 | 0.032 | 0.050 | 0.018 | 0.004 | 10 |
|  |  | 3 | 5 | 13% | 0.049 | 0.049 | 0.037 | 0.058 | 0.020 | 0.005 | 9 |
|  |  | 4 | 13 | 33% | 0.060 | 0.059 | 0.050 | 0.071 | 0.021 | 0.006 | 9 |
|  |  | 5 | 30 | 75% | 0.086 | 0.087 | 0.071 | 0.096 | 0.024 | 0.005 | 6 |
|  |  | 6 | 39 | 98% | 0.119 | 0.119 | 0.107 | 0.134 | 0.028 | 0.005 | 4 |
| 5 | A5 | 1 | 4 | 10% | 0.021 | 0.021 | 0.005 | 0.029 | 0.023 | 0.005 | 23 |
|  |  | 2 | 14 | 35% | 0.041 | 0.041 | 0.029 | 0.055 | 0.026 | 0.006 | 15 |
|  |  | 3 | 23 | 58% | 0.050 | 0.050 | 0.040 | 0.064 | 0.023 | 0.005 | 11 |
|  |  | 4 | 24 | 60% | 0.061 | 0.061 | 0.048 | 0.071 | 0.023 | 0.005 | 8 |
|  |  | 5 | 39 | 98% | 0.085 | 0.086 | 0.075 | 0.093 | 0.018 | 0.005 | 6 |
|  |  | 6 | 40 | 100% | 0.120 | 0.121 | 0.108 | 0.131 | 0.022 | 0.005 | 4 |
|  | B5 | 1 | 3 | 8% | 0.019 | 0.019 | 0.011 | 0.029 | 0.018 | 0.005 | 25 |
|  |  | 2 | 8 | 20% | 0.040 | 0.040 | 0.028 | 0.049 | 0.020 | 0.005 | 12 |
|  |  | 3 | 15 | 38% | 0.050 | 0.049 | 0.039 | 0.066 | 0.027 | 0.005 | 11 |
|  |  | 4 | 17 | 43% | 0.060 | 0.059 | 0.051 | 0.070 | 0.019 | 0.005 | 9 |
|  |  | 5 | 34 | 85% | 0.085 | 0.085 | 0.076 | 0.093 | 0.017 | 0.004 | 4 |
|  |  | 6 | 37 | 93% | 0.120 | 0.119 | 0.098 | 0.133 | 0.035 | 0.006 | 5 |

Abbreviations: Std, standard deviation; CV, coefficient of variation.
<sup>a</sup>Hit rate: the proportion of 40 replicates per level that are test positive.

**Figure S1.**
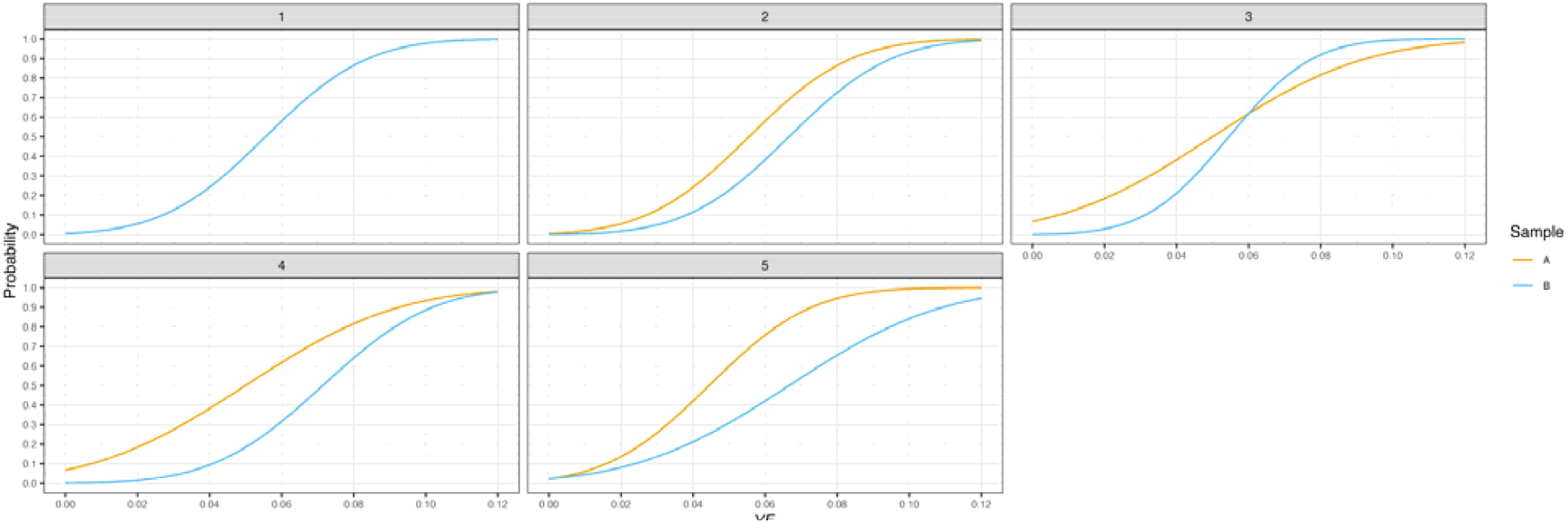
Theoretical probit regressions derived using parameters used in simulated scenarios. The orange line denotes the surrogate sample A, and the blue line denotes the patient sample B.

## Footnotes

a Centering by the mean of the VFs across replicates spanning the six dilution levels of the two samples.

b If the assumptions of probit regression are not fully met, some corrective steps may help reduce deviations. For instance, if the errors are not normally distributed, an arcsine transformation on VF may make the errors normally distributed and stabilize the variances across the dilution levels.

c A logit regression (with logistic distribution as the link function) may be used instead. Since CLSI EP17 (8) has detailed the use of probit regression in determining LOD, probit regression is used throughout this work.

d Surrogate and patient sample data may not be poolable due to various reasons, such as substantially different experimental conditions, different bioinformatic or analytical pipelines, or strong lab batch effects.

